# A comparative study of infection and mortality in COVID-19 across countries: A scaling analysis

**DOI:** 10.1101/2020.10.13.20212175

**Authors:** Ranjan Das, Akmal Hossain, Sayak Mandal, Debasmita Pariari, Rohit Kumar Rohj, Swapnil Shukla, Rahul Mahavir Varma, D. D. Sarma

**Author notes:** These authors contributed equally and their names appear in alphabetical order (Ranjan Das, Akmal Hossain, Sayak Mandal, Debasmita Pariari, Rohit Kumar Rohj, Swapnil Shukla, Rahul Mahavir Varma).

## Abstract

Analysing infection and mortality data for COVID-19 as a function of days for 54 countries across all continents, we show that there is a simple scaling behaviour connecting these two quantities for any given nation when the data is segmented over few ranges of dates covering the most rapid spread of the pandemic and the recovery, wherever achieved. This scaling is described by two parameters, one representing a shift along the time axis and the other is a normalisation factor, providing a reliable definition of the mortality rate for each country in a given period. The number of segments for any country required in our analyses turns out to be surprisingly few with as many as 16 out of 54 countries being described by a single segment and no country requiring more than three segments. Estimates of the shift and mortality for these 54 countries in different periods show large spreads ranging over 0-16 days and 0.45-19.96%, respectively. Shift and mortality are found to be inversely correlated. Analyses of number of tests carried out for detecting COVID-19 and the number of infections detected due to such tests suggest that an effective way to increase the shift, and therefore, decrease mortality, is to increase number of tests per infection detected. This points to the need of a dynamic management of testing that should accelerate with the rise of the pandemic; it also suggests a basis for adjusting variations in the testing patterns in different geographical locations within a given country.

---

COVID-19 does not need an introduction; the pandemic has made that superfluous. Unarguably, there is no other scientific term that has ever been more widely known to the world population. Naturally, this has led to an immense effort within the global scientific community to understand its possible nature, trajectory, consequences, remediation and treatments, leaving aside more contentious aspects like its probable origin, relative merits, based on their efficacies, of various national efforts, and the balancing acts between the management of the pandemic and the financial health. These efforts, as a consequence of the sense of urgency involved, have led to a proliferation of various analyses, speculations, hypothesis, and claims appearing in every form of communication available to us. With this background, we need to justify why we bring out this report when we seem to have a glut of information already swamping our sensitivities and sensibilities.

It is believed that the number of people infected with COVID-19 is much larger than the number of affected people with a large proportion of infected ones remaining asymptomatic. Even among the group affected with perceptible symptoms, it is only a small fraction that develops serious health complications, including mortality in extreme cases. The presently known crisis faced by the humanity arises from this small fraction with the total number of infected or the number with milder symptoms being essentially ignored. It has even been said that the number of infected being large is in fact desirable in terms of attaining herd immunity quickly, as long as the mortality rate can be kept low. In absence of any viable treatment at present, the only way to control the mortality rate is through early detection and proper medical care and support, giving the body the best chance and the longest time to tackle the virus. While medical sciences grapple with the all-important issues of understanding various aspects pertaining to the interaction of the SARS-CoV-2 with the human system and drug and vaccine candidates at the biological levels, it is instructive to see if the vast amount of information available on the mortality rates from different countries provide us any generalisation despite the extreme diversity in national mortality rates. In fact, one may turn around this observation and ask the question that has often been asked about the origin of this tremendous spread of mortality rates between different nations. Understandably, this is a complex question that is necessarily dependent on many factors, some possibly even unknown to us at present. So, we reduce the question to a more manageable one by asking if we can identify some of the factors that may have an influence on the mortality rate specifically and on the trajectory of this pandemic in general.

The data that are readily available for analyses from most countries are the cumulative and daily numbers of infections detected and deaths registered related to this pandemic. However, there are considerable disparities and uncertainties related to the definitions applied by each nation to build up these databases. One specific and easily identifiable example of such uncertainties can be seen in terms of a sudden spike in the infection or death count that is evidently far beyond any statistical fluctuations, as illustrated in Fig. 1(a) for the daily death counts reported by India ^1^ with red dots. This arises from an attempt to correct daily death counts due to missing reports over a period by accounting for all such cases in a single day, ^2^ giving rise to such an unreal spike in the death count for that single day. This naturally leads to a discontinuity in the cumulative death count on that particular day, as shown in Fig. 1(b) with red dots. In order to deal with this specific type of inaccurate data from vitiating the analysis, we distribute the excess count of deaths (or infections, as the case may be) on that particular day over each day of the preceding period in a way proportional to the already reported death (or infection) count of that day; this amounts to assuming that a fixed percentage of deaths (or infections) was uniformly missed each day leading to the total number of missed counts being corrected. We illustrate this procedure by applying it to data in Fig. 1(a) and (b) where the original data with the spike in the count appear as red dots and the derived data, with the count in the spike distributed over all preceding days proportionately, as open blue circles. Clearly, the two data sets for daily death counts are almost identical everywhere except for the unreal spike on the single day being absent in the resulting data set. Likewise, the two data sets for the cumulative number of deaths are essentially identical except in the vicinity of the anomalous day with the corrected data presenting a smooth interpolation of the discontinuous change in the original data set. Such corrections are required only for a limited number of cases, namely for China, South Africa, Chile, Portugal, Hungary, India, and USA.

**Figure 1:**
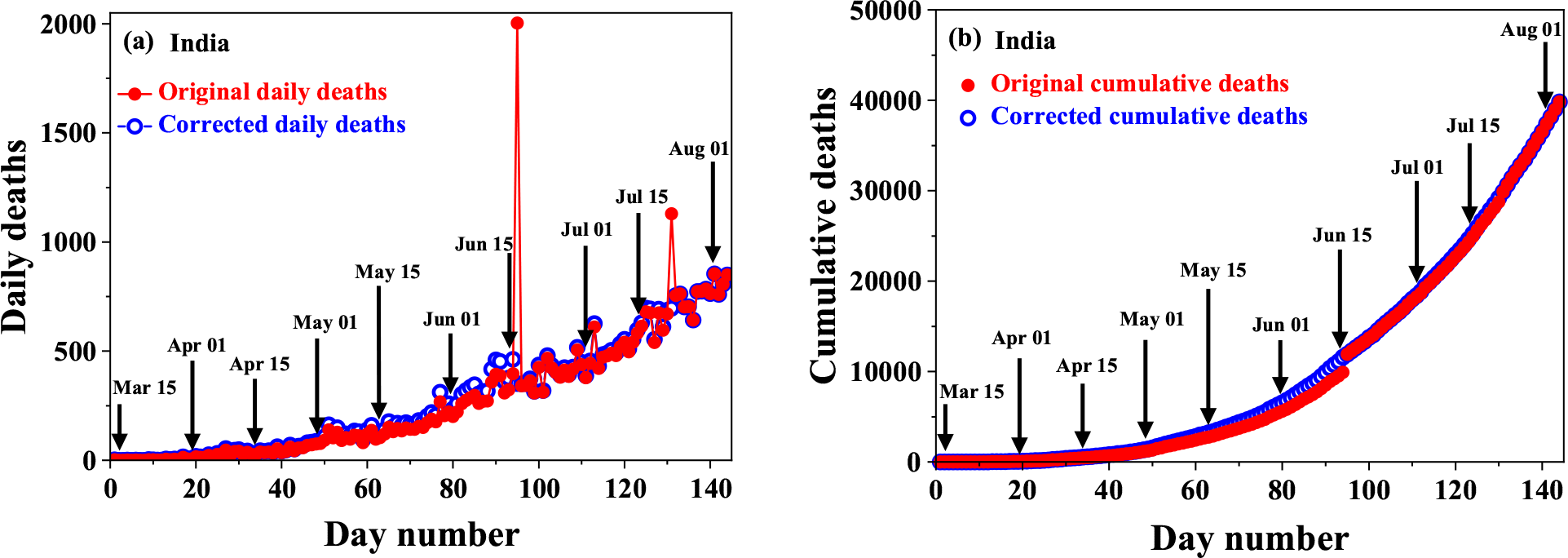
(a) Daily death count as reported with two clearly identifiable spikes at day numbers *95 (16 Jun)* and *131 (22 Jul)* shown with red dots. The derived data after distributing the excess counts of these two days over the preceding periods, as discussed in the text, are shown with open blue circles. (b) Cumulative death count (red dots) as reported, exhibiting discontinuous jumps corresponding to the spikes in the daily counts are shown together with the derived cumulative death count (open blue circles) as a function of the day after distributing the anomalous death count in the spikes over the preceding periods.

While the above procedure appears reasonable for such recognisable anomalies in the death count, there are many other uncertainties in data reporting that are impossible to account for. Thus, any conclusion based on analyses of such uncertain databases must be robust against such diversities of definitions and accounting of the numbers of infected and deaths. The hope is that analyses of extensive data from many nations will make the conclusions statistically useful in showing us underlying trends, if any exist. If the mortality would depend only on the virulence of the virus vis-á-vis the immunity level of the population of a nation, in absence of any well-established medical intervention, the mortality rate is fixed for the nation, independent of any national policy and measures to contain the infection, such efforts only influencing the spread and consequently, the number of infections and deaths, but not the ratio of the two that is defined as the mortality. An estimate of the mortality for any nation is easily obtained by taking the ratio of the total cumulative numbers of deaths and infections on any given date. This ratio is readily seen as the lower bound of the mortality, since on the date of the evaluation, there are a certain number of active infections with an inherent mortality rate that will increase the total number of deaths with time even if there is no other fresh infection detected.

Since the detection of the infection generally precedes the event of a death in a patient, there is often a lag between the infection and the mortality in any given case of mortality that we call the shift, measuring simply the number of days between the detection and the death. While the mortality rate may not be affected by various national measures, as argued above, clearly this shift depends critically on such measures, for example a widespread testing program, leading to an early detection and enhancing the shift. While this shift is precisely definable for each case of mortality, a-priori, it is not obvious that there will be any recognisable measure of this shift for a nation, since in reality there are distributions of every relevant aspects controlling the infection/death correlation within any given nation, such as the stage of the detection, strategies of testing, virulence of the infection in individuals, definition and recording of the deaths due to COVID-19, immunity level of individuals, availability of the health-care system and other presently imponderable factors. Different geographical locations within a country may also have very different distributions of these aspects that are expected to have an impact on any statistical measure of the shift between the detection of the infection and the death. Even more important is to acknowledge that some of these factors even within any single nation may change with time as the society responds to the surging pandemic with various social, political, and medical interventions. Analysing the infection and mortality data from 54 countries seriously impacted by this pandemic across all continents, we surprisingly find that it is indeed possible to have well-defined values of Shifts (*S*) for every nation despite these variabilities. A consequence of this ability to estimate *S* reliably is that it allows us a more reliable estimate of the mortality rate (*M*) with the identification of the relevant date of the infection to be related to the mortality on a certain date, with these two dates being separated by *S* in the national statistics. Not only we find large spreads in the obtained *S* and *M* across the countries analysed, there is clearly an inverse correlation between the two, despite our naïve arguments earlier to suggest that *M* should not depend on any national measures. We note that the knowledge of these two scaling parameters for a given country allows it to anticipate the demand on the health-care system from the measured number of infections on any given day, a longer shift giving the nation more time to respond and a lower mortality rate, of course, placing less demand on the system. Finally, we analyse the detection/test data of all these countries to show how *S* can be influenced for an effective management strategy.

We illustrate the methodology of our analysis with the example of Switzerland, whose progressive cumulative infection number is plotted using the left axis in Fig. 2(a) with red open circles against the Day Number with the Day Number = 1 being March 12, 2020; we have also marked the beginning and the middle of every month in the plot to give a better idea of the transition of calendar dates through the plot-period. In the same plot we have also shown the progressive cumulative number of deaths (black open circles) using the right axis over the same period. The relative scales of the left and the right y-axes have been adjusted to make the two plots appear normalised at the end point to make obvious the shift between the two plots along the time axis. Denoting the dependencies of the number of cumulative infections (*N*_*i*_) and cumulative deaths or mortality (*N*_*m*_) on the day number (*n*_*d*_) as *N*_*i*_(*n*_*d*_) and *N*_*m*_(*n*_*d*_), we obtain values of two parameters, Factor (*F*) and Shift (*S*), such that the scaled function,

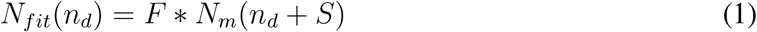

provides the best description of *N*_*i*_(*n*_*d*_) in the least-squared-error sense, making

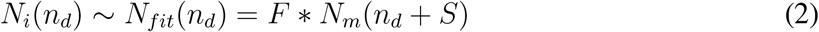

**Figure 2:**
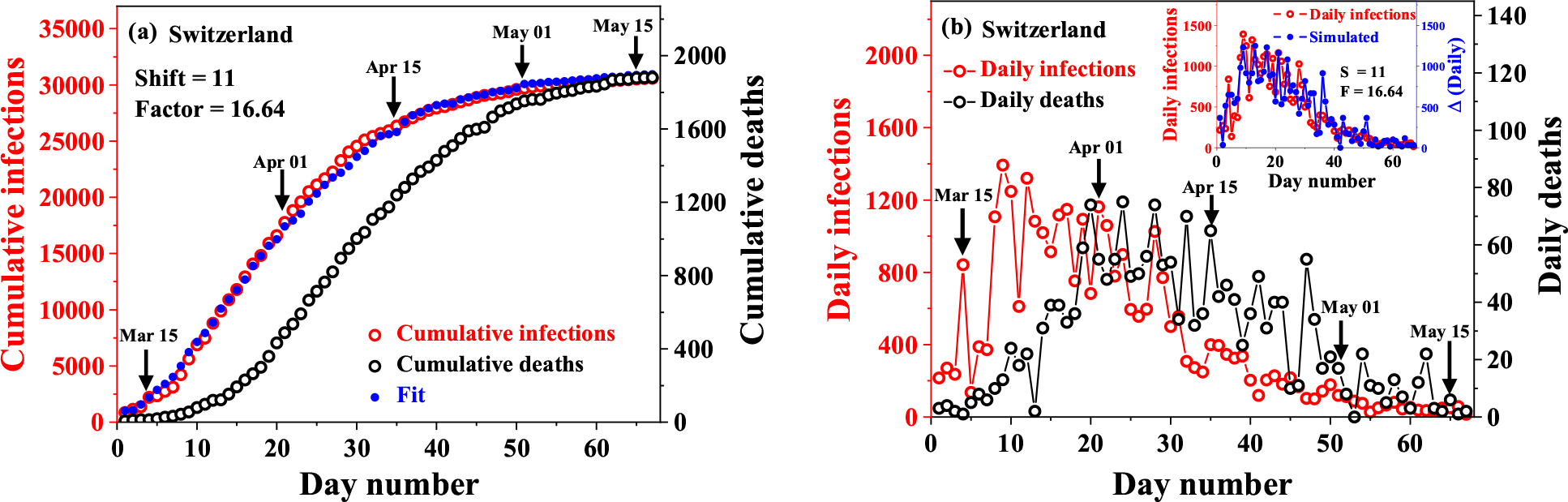
(a) Cumulative number of infections (red open circles) and deaths (black open circles) for Switzerland as a function of the day with day 1 = March 12, 2020. The cumulative number of deaths shifted by 11 days and multiplied by the factor (16.64) is shown with blue dots to illustrate the scaling behaviour. (b) The number of daily infections and deaths over the same period as in panel (a). The inset shows the result (Δ) of shifting the number of daily deaths by 11 days and multiplying by 16.64, as determined from the scaling of the cumulative numbers of infections and deaths in panel (a) leads to a good correspondence for the daily counts of these two quantities as well.

In other words, we find the shift, *S*, in the number of days and the corresponding multiplicative factor, *F*, that when applied to the cumulative mortality numbers give the best approximation to the observed cumulative number of infections. Since percentage mortality is defined as the number of deaths for every 100 infections, the factor, *F*, directly provides the mortality rate as 100*/F* and we report throughout this paper the number for the percentage mortality rate estimated via this method. In the specific case of Switzerland, this least-squared-error approach provides us with a shift and mortality rate of 11 days and 6.01% and the resulting scaled cumulative function, *N*_*fit*_(*n*_*d*_), is shown with blue dots in Fig. 2(a); the comparison of the measured infection data, *N*_*i*_(*n*_*d*_), plotted with red open circles and the fitted or scaled data, *N*_*fit*_(*n*_*d*_), plotted with blue dots using the same common left axis makes it evident that this scaling works well for Switzerland with just a single value of Shift describing the entire period March 12-May 17, 2020 spanning more than two months that we analysed.

We note that the incidence of infections and deaths in Switzerland beyond this period dropped to relatively small numbers, making it statistically insignificant. This is illustrated in Fig. 2(b) where we plot daily infections and deaths, in contrast to the corresponding cumulative numbers plotted in Fig. 2(a). In Fig. 2(b), the red open circles, referred to the left axis and representing the number of daily infections reported, do not appear to be in any obvious relationship with the black open circles, referred to the right axis and representing the number of daily deaths reported, in the main panel. However, when we use the same scaling factor and shift on the number of daily deaths, shown in blue dots in the inset to Fig. 2(b), and compare with the reported daily infections (in red), the correspondence becomes obvious, establishing the scaling behaviour extracted from the analysis of the cumulative numbers and illustrating our methodology of deriving the Shift (*S*) and the Factor (*F*) for each country.

We found that the above described approach works very well for 16 of the 54 countries that we analysed. For the remaining 38 countries, it proved impossible to find such a single scaling set to map the number of deaths on to the number of infections in the cumulative or the daily counts over the entire period under consideration. We illustrate this difficulty in Fig. 3(a) with the example of data available for Japan over the period March 5-May 30, 2020. The single segment analysis, successful for Switzerland (Fig. 2), was applied to the case of Japan and the least-squared-error procedure yielded Shift = 15 and Factor = 19.37. When the cumulative death number (black open circles) were scaled with this set of scaling parameters, it results in plot shown with blue dots as the best single-segment fit to the cumulative infections reported (red open circles), illustrating the failure of the single-segment analysis.

**Figure 3:**
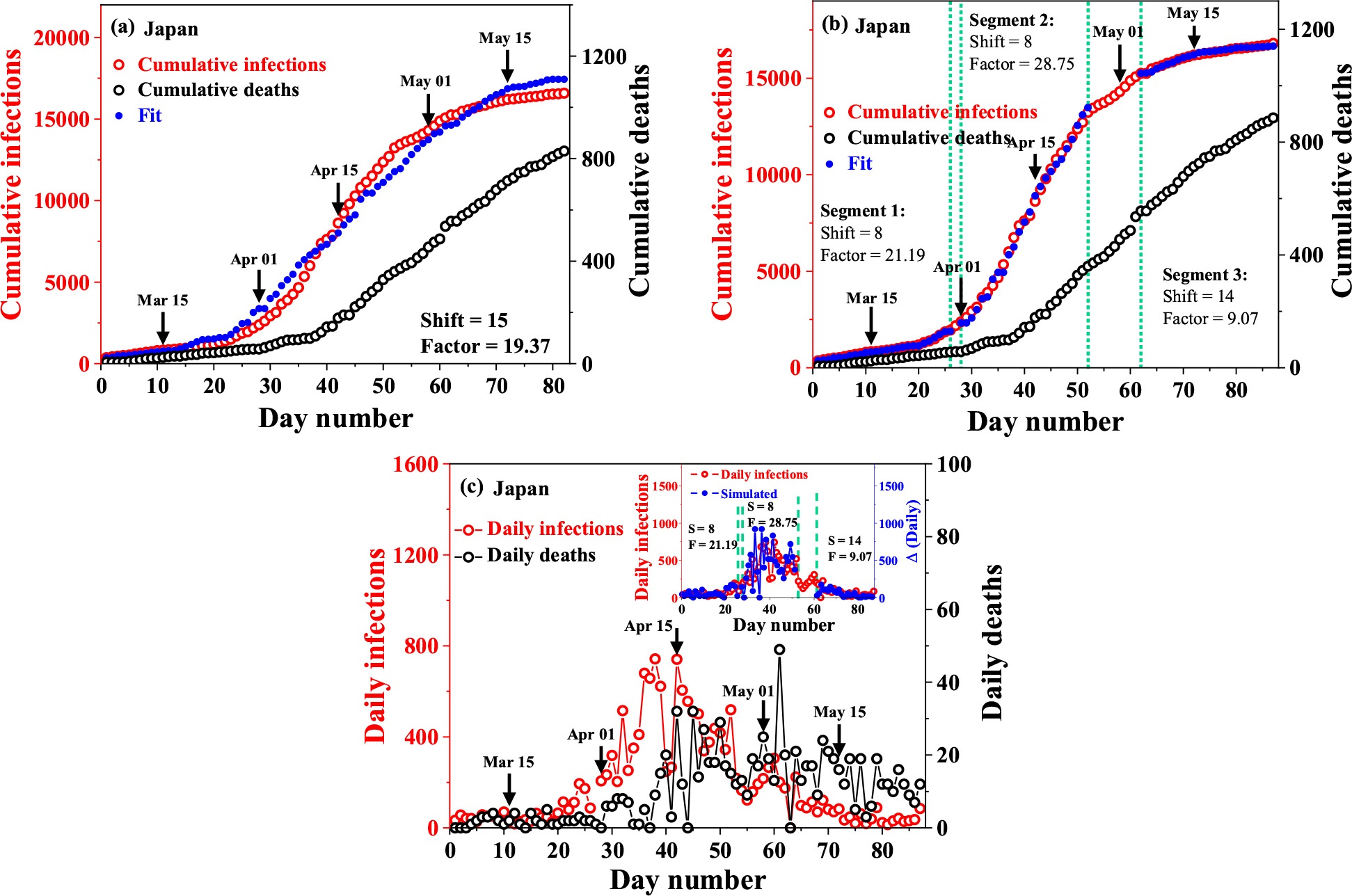
(a) Cumulative number of infections (red open circles) and deaths (black open circles) for Japan as a function of the day with day 1 = March 5, 2020. The blue dots represent the single segment scaling (see text) with the optimal Shift = 15 and Factor = 19.37, providing a poor description of the measured cumulative infections (red open circles). (b) Same as in panel (a), but the scaling analysis is carried out in three segments as marked with independently determined optimal Shift and Factor for each segment. The three-segment scaled plot is shown with blue dots, exhibiting a good description of the measured cumulative infections (red open circles). (c) The number of daily infections and deaths over the same period as in panels (a) and (b). The inset shows the result (Δ) of scaling the number of daily deaths (blue dots) in each of the three segments marked in panel (b) by optimal shifts and multiplying by optimal factors, as determined from the three-segment scaling of the cumulative numbers of infections and deaths in panel (b), leading to a good correspondence for the daily counts of the reported number of infections (red open circles) and the scaled daily deaths.

This prompted us to carry out multi-segment analysis, increasing the number of segments systematically to provide a good description of the reported cumulative infection by the scaled cumulative death with the least number of segments. We found that the case of Japan required a minimum of three segments with each segment independently scaled via the least squared-error approach, to arrive at a good agreement between the reported data and the scaled plot, as shown in Fig. 3(b), where we have also marked the three segments with vertical dashed lines and indicated the Shift and the Factor for each segment. The consistency of the derived shifts and factors from this 3-segment analysis with the data of daily infections and deaths is illustrated in Fig. 3(c), where the main frame shows the plots of the daily infections (red open circles) and daily death counts (black open circles) with no evident similarities between the two. In the inset we show the scaled daily death counts (blue dots) in the three separate segments with the corresponding shifts and factors, as shown in Fig. 3(b), in comparison with the daily count of infections (red open circles), illustrating a very good agreement between the two.^1^

Out of the 38 countries requiring such multi-segment analyses, 14 required 2-segment analyses and 24 required 3-segment analyses. We have summarised all necessary information against each of the 54 countries analysed by us in Table 1. This table also illustrates that even in such multi-segment analysis, it is possible to identify one specific segment as the dominant one with the maximum number of infections reported during that period. For example, in the case of Japan shown in Fig. 3, the three segments, defined by the date periods of March 5-March 30 (Segment 1), April 1-April 25 (Segment 2), and May 5-May 30 (Segment 3), had 1622, 11053, and 1726, respectively, reported cases of infections (see Table 1), clearly establishing Segment 2 as the dominant segment. We use this definition of a dominant segment for each country that requires a multi-segment analysis, the remaining segment(s) being termed as minor.

**Table 1:**
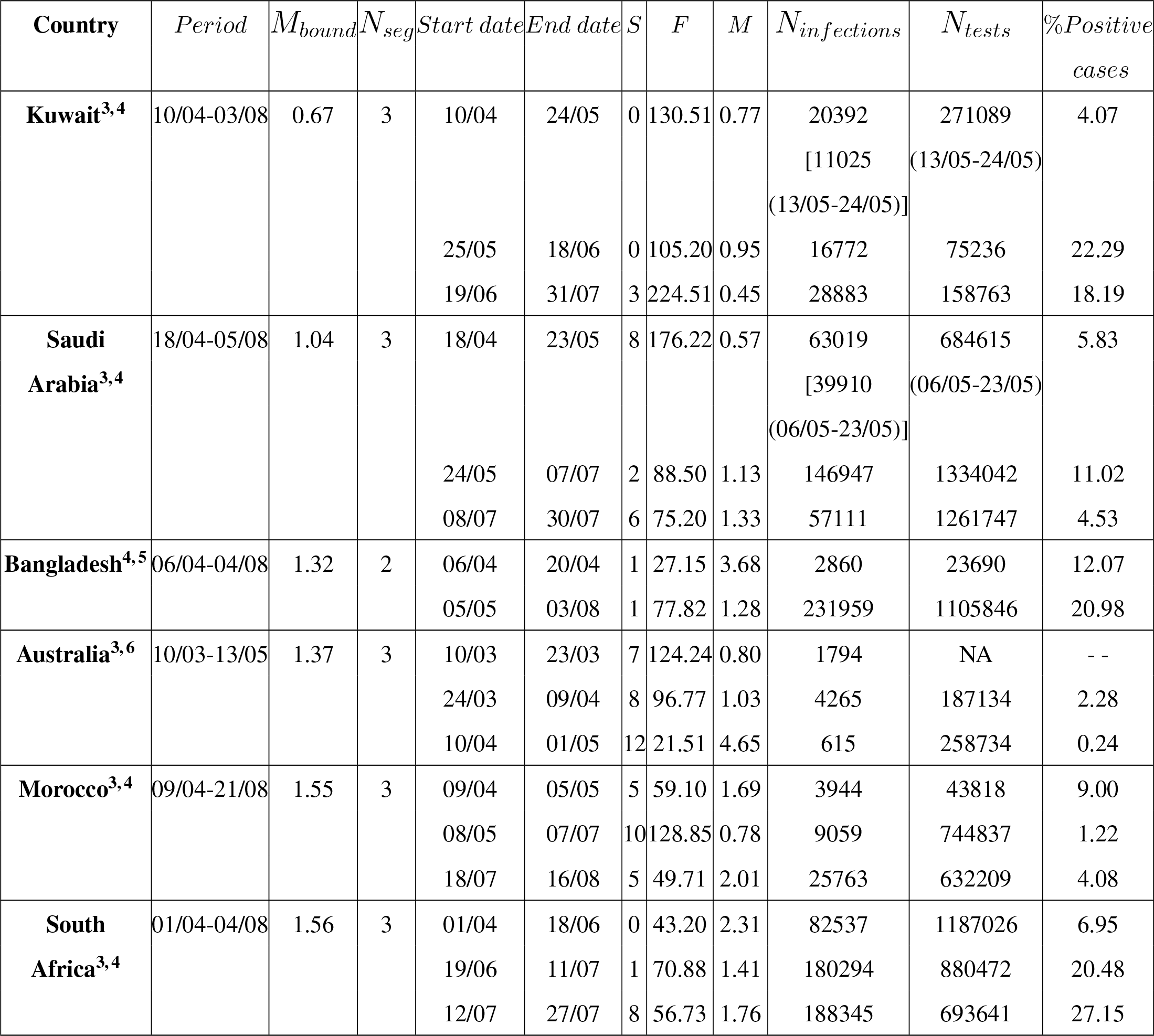
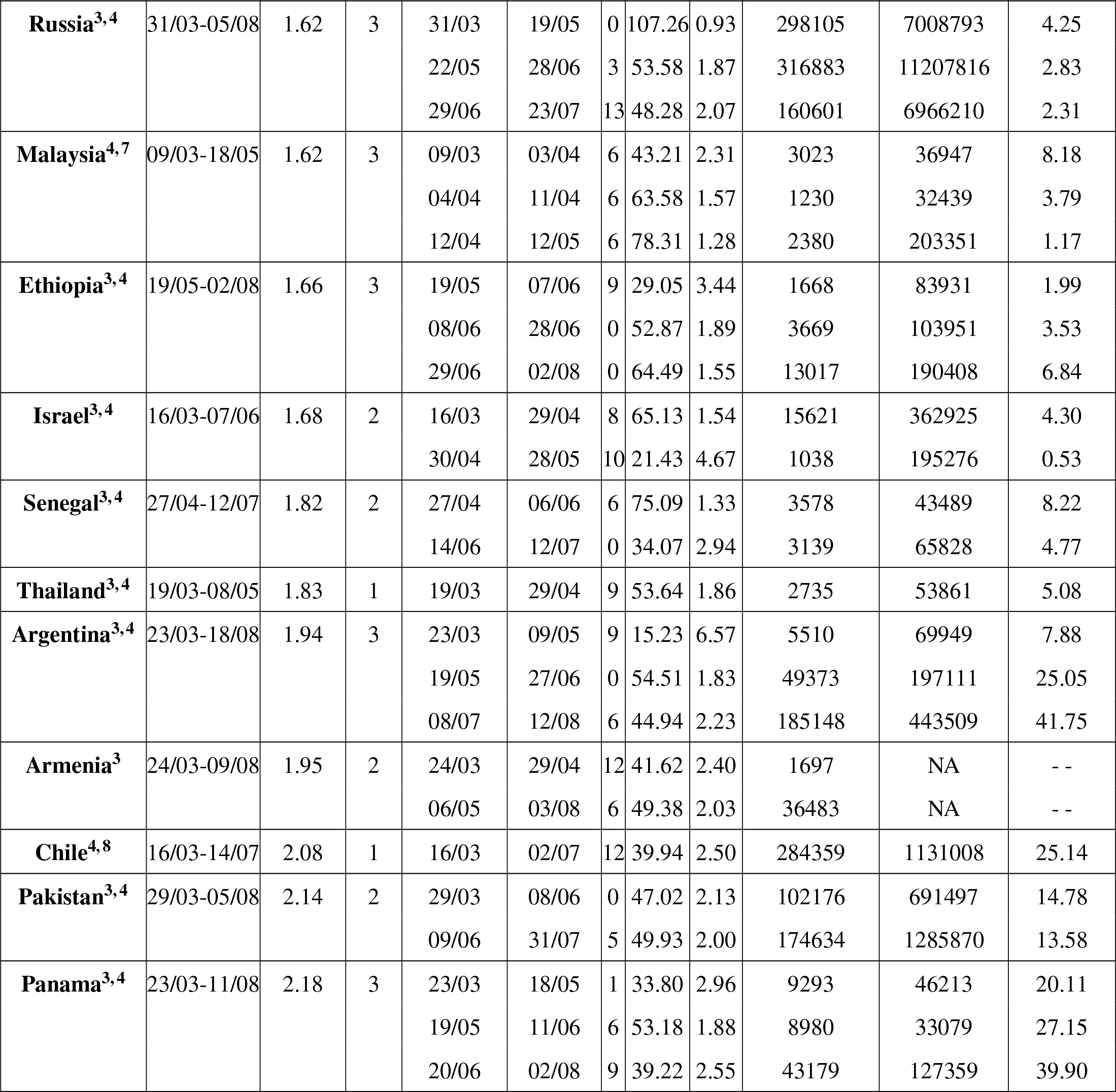
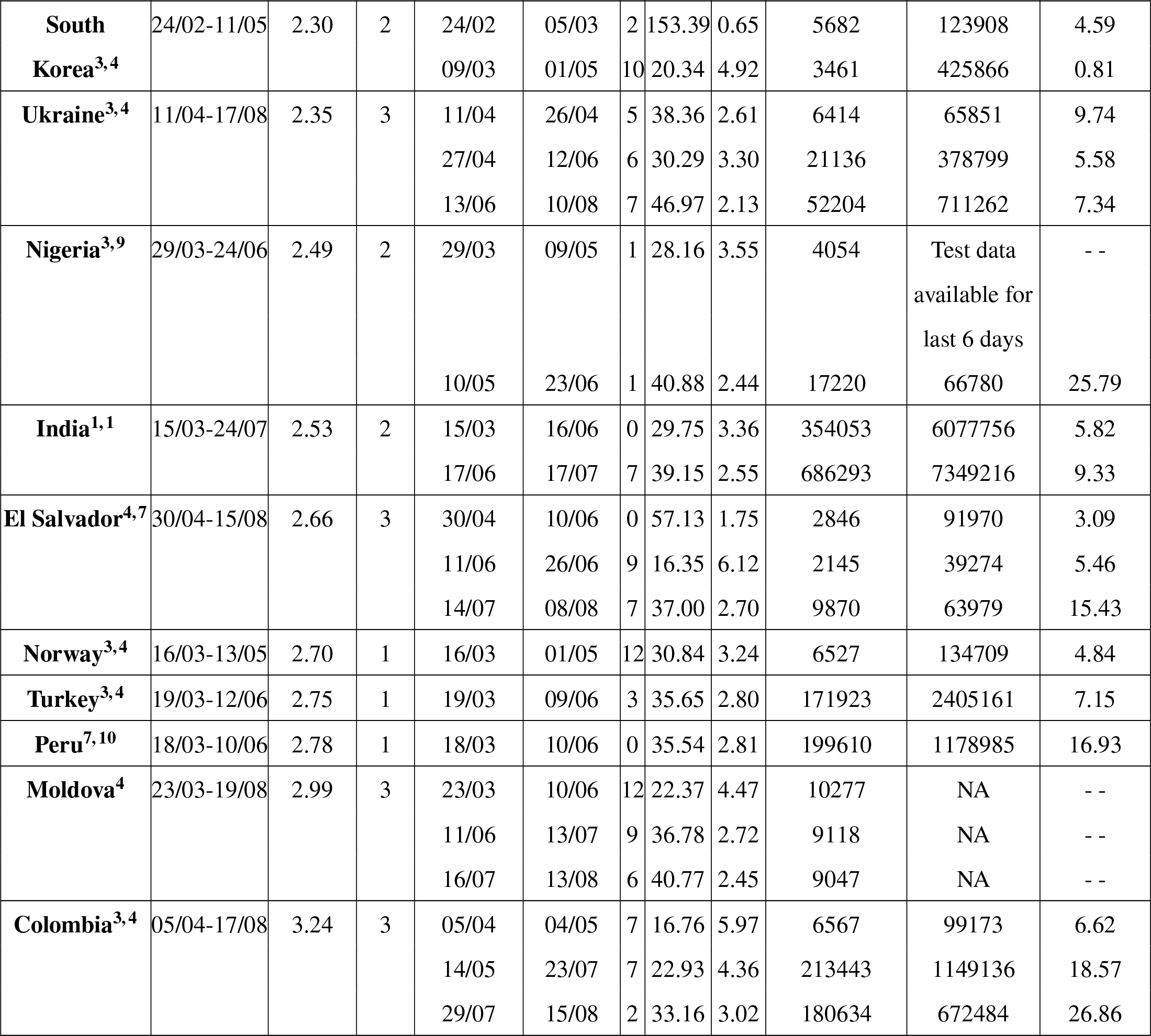
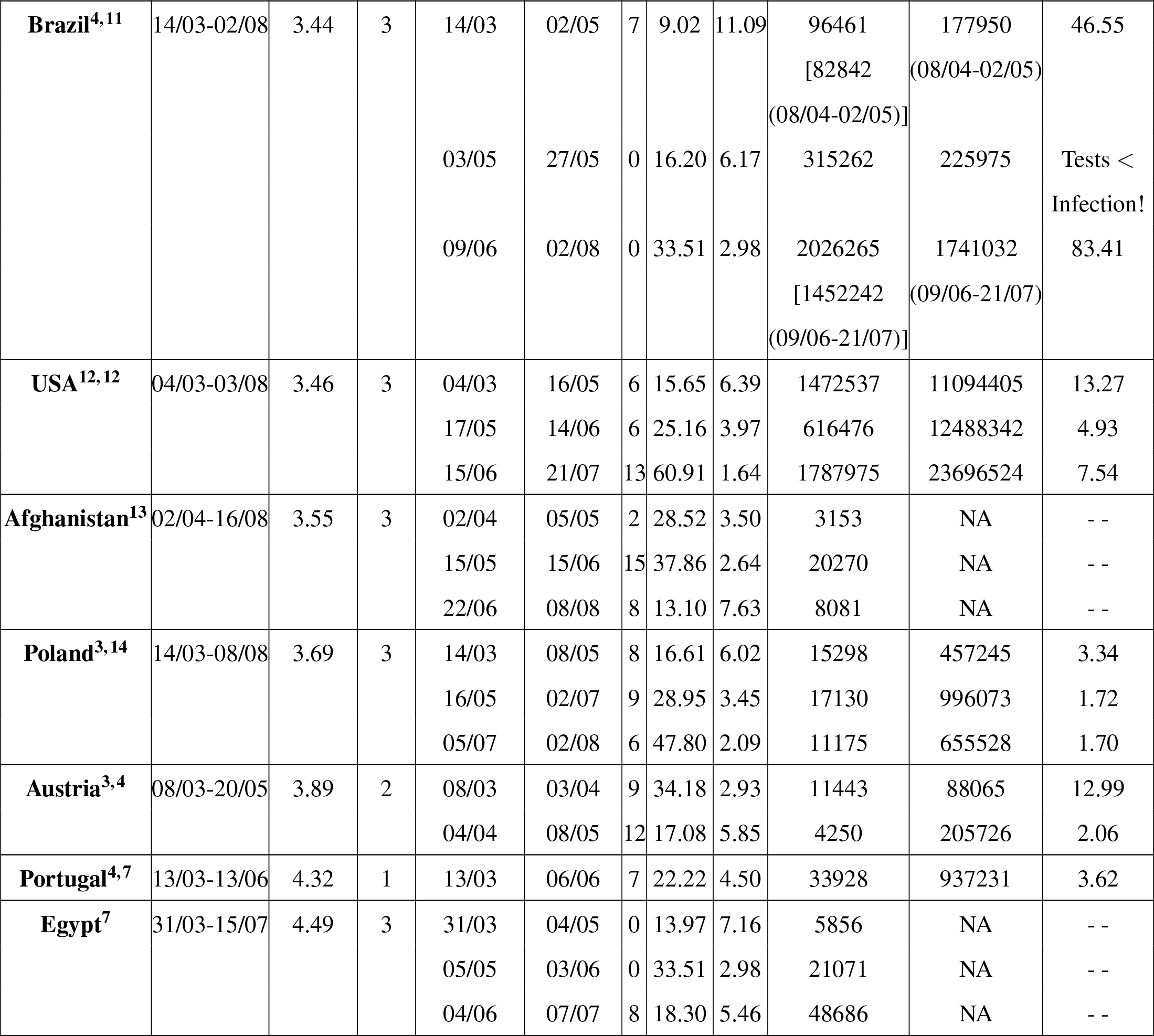
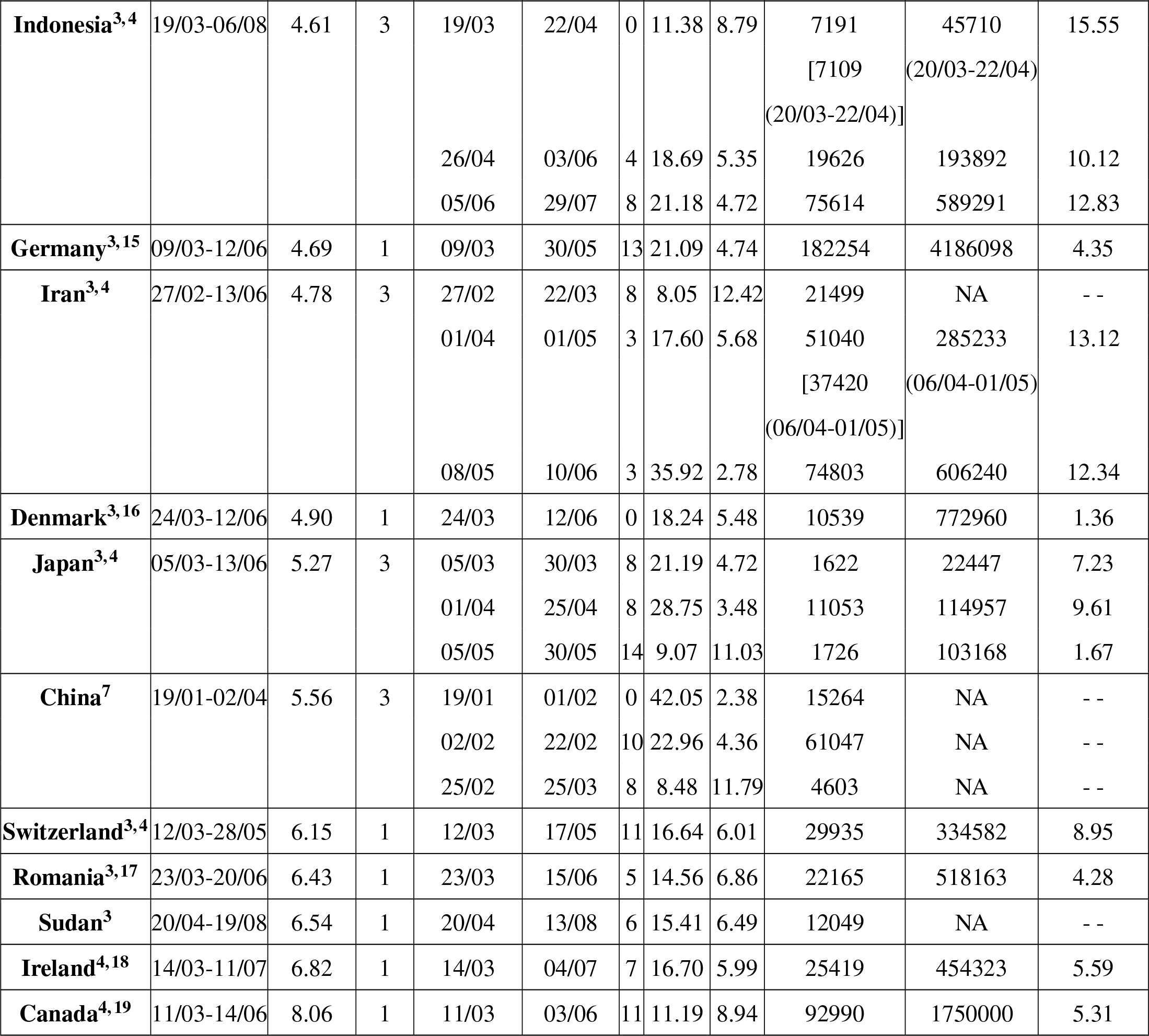
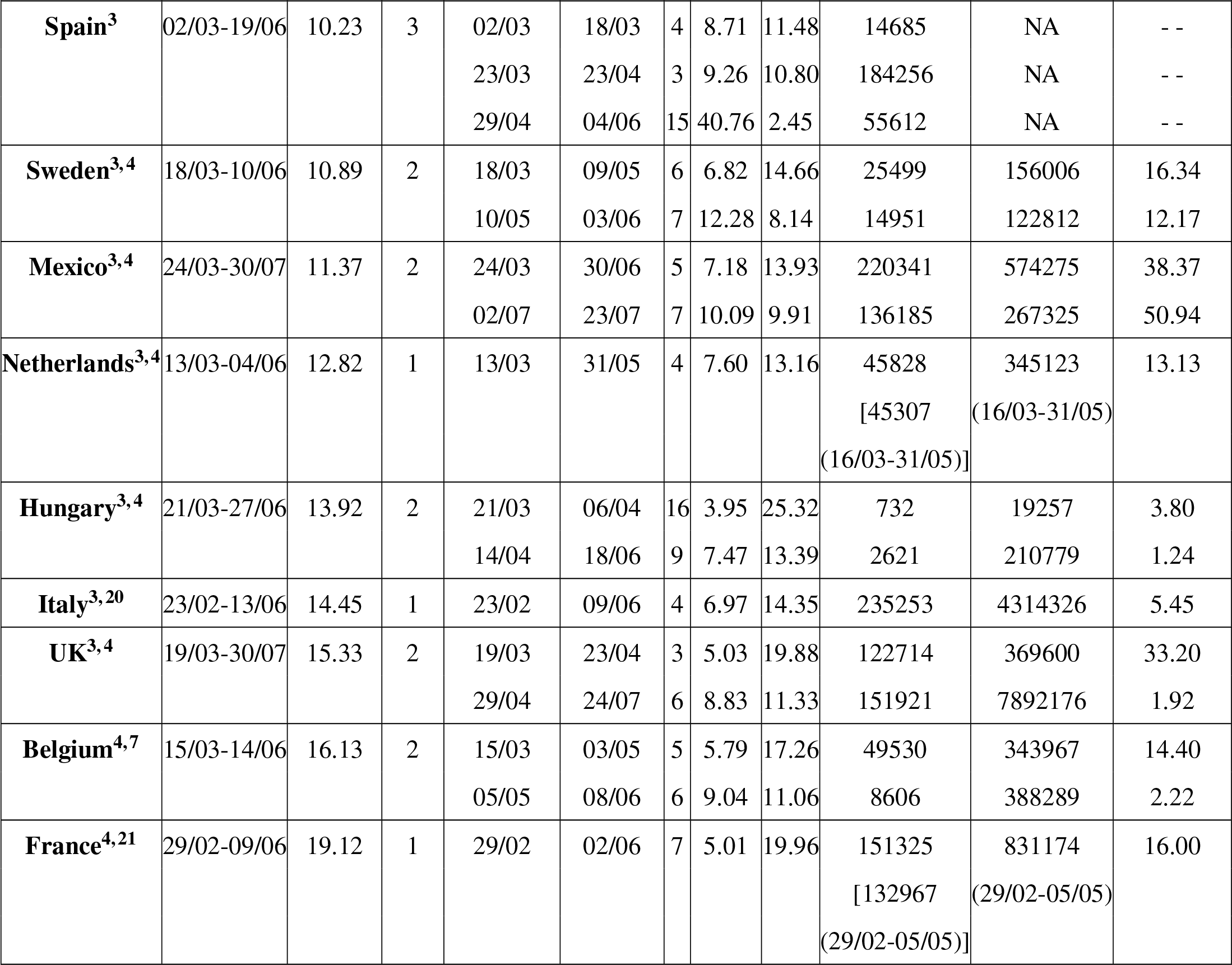
All details for each country analysed in this work along with the analysis results. Explanations of abbreviations are given at the end of this table. The two citiatons shown as superscripts separated by a comma after each country indicates the source of the data, one of them is pointing to the infection and mortality data, and another is pointing to the testing data.

As already mentioned, the calculated parameter, Factor (*F*), is related to the mortality rate (*M*), as *M* = 100*/F* ; these estimates of the mortality rates for all countries and different segments are shown in Table 1 in addition to the values of the Shift, *S*, and *F*. We note that another definition of the mortality rate for any given country can be the total number of deaths divided by the total number of infections on the last date included in our analysis; we already pointed out that this defines a *lower bound* of the mortality. We explore how the estimate of the mortality rate as 100*/F* relates to this lower bound of the mortality rate for various countries by plotting one against the other in Fig. 4. The diagonal dashed line is the 45^*°*^ line, indicating the trajectory of data points if the presently defined %mortality (= 100*/F*) is equal to the lower bound. Thus, one expects that all data points to be either on this dashed line or above it since there cannot be any %mortality lower than the bound. The red dots are for countries with single-segment analysis, showing a very good correlation with the lower %mortality bound. While the %mortality is found to be spread over a wide range, from 1.86% for Thailand to 19.96% for France, all the red dots appear close to the dashed line, either being virtually on it or approaching it from above. Since, in the long run, %mortality must be equal to the lower bound, a sizable gap between the two for the period of analysis suggests that these countries are still far from reaching the end of the pandemic on the last date included in our analysis. Countries which have flattened the curve, as it is called, reaching the low incidence of infection and mortality by the end date of our analysis show that the %mortality estimated by the scaling is almost equal to the lower bound with the red dots of these countries appearing on the dashed line.

**Figure 4:**
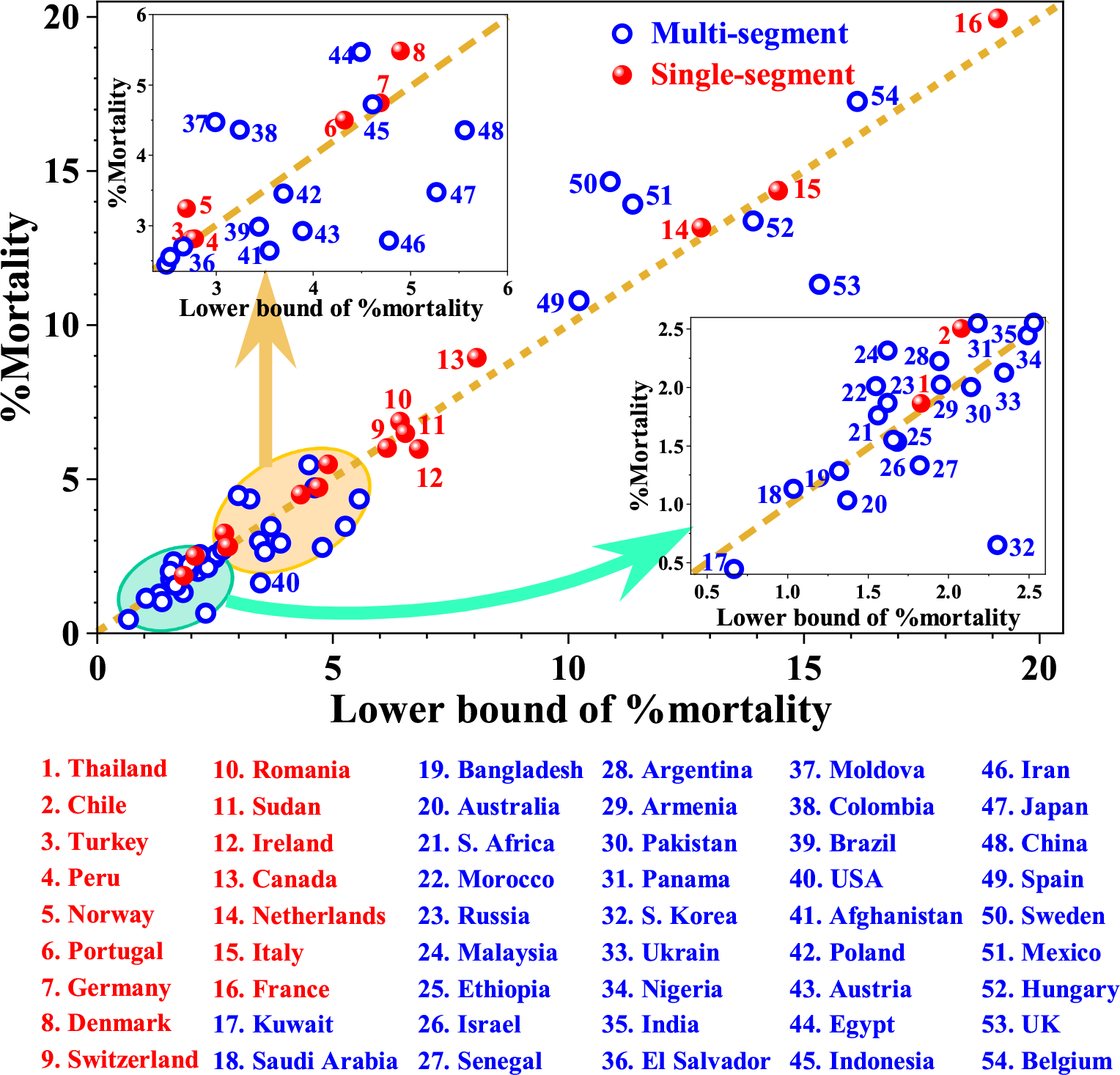
Plot of %Mortality estimated from the scaling analysis vs. the lower bound of %mortality estimated from the total numbers of mortality and infections for each country.

We have also plotted *M* of the dominant segment for each country requiring a multi-segment analysis in Fig. 4 with a blue open circle. It should be noted that in this case, *M* is not limited by the lower bound, in contrast to the single segment analyses, since the lower bound refers to the data collected over, not only the dominant, but all segments. Thus, it is not surprising to find some of the %Mortality of the dominant segment being lower than the lower bound for a few of the countries; it suggests that the country did relatively well in keeping the mortality rate low during the incidence of the majority of the infections or alternately, had a high rate of %Mortality during the minority segment(s). The primary examples of this behaviour are UK, Iran, South Korea, USA, and Japan (see Fig. 4 and Table 1). The reverse case is found to be true for Sweden and Mexico with a high *M* for the dominant segment. In any case, a clear correlation between *M* and the lower bound of the mortality from the total deaths and infections is established by Fig. 4, suggesting the relevance of the parameter *M* obtained from our scaling analysis as a definition of the mortality rate for each country over the specified time interval (Table 1).

The other parameter, Shift (*S*), measures the typical number of days between the detection of the infection and the occurrence of the mortality for a country during the time segment defined in Table 1. Interestingly, this scaling parameter also shows a huge variation between countries with its value ranging from 0 to 16. While different countries may have different protocols of registering the mortality date, making comparisons between countries somewhat difficult, it is clear that given any protocol for a country, larger the *S* is more time is available for the person to receive medical support and critical care, increasing the chances of survival. Therefore, it is reasonable to anticipate an inverse correlation between the shift (*S*) and the %mortality (*M*) if other factors remain invariant. With this in mind, we have plotted the %Mortality (red dots and blue open circles) vs. the Shift for every country in Fig. 5 using the results of the dominant segments (shown with blue open circles) for countries with multi-segment analysis. In a few cases, minor segments have comparable number of infections as in the dominant segment, making the identification of the dominant segment somewhat arbitrary for such countries. Specifically, we find that each of UK, USA, Russia, Senegal, and South Africa have a segment with the number of infections within 80% of that in the dominant segment of that country. We include results of analysis of such segments, referred to as significant-segments, shown with stars in Fig. 5. Evidently, Fig. 5 shows some interesting trends, being restricted to only certain regions (highlighted by the light green shade) of the Shift-%Mortality phase space. The highlighted region shows two distinct parts of the phase space being populated, one for the low %Mortality (⩽ 3.0) region with a small slope suggesting relative insensitivity of the %Mortality on the Shift. Intriguingly, all developing countries, with the exception of Egypt, Romania and Mexico, appear in this part of the phase space. It is also interesting to find that almost all countries, except Egypt, with hot/warm weather with temperatures reaching beyond 30 C during the period under consideration appear in this group of low mortalities and varying shifts. However, this group with low mortalities also includes Russia, South Africa, South Korea, Australia, and Austria with relatively colder climates during period being analysed here, suggesting that along with many other factors, the temperature may also be playing a significant role in controlling the mortality rate in a given nation. While the negligible inclination of this shaded region encompassing the group with a low mortality rate (⩽ 3%) suggesting %Mortality is not influenced significantly by the Shift, the highlighted region with a prominent inclination encompassing the remaining countries with %Mortality varying between ∼1 and 20 and Shift varying between 3 and 15 defines a clear correlation between these two parameters with a rapidly decreasing mortality rate with an increasing shift.

**Figure 5:**
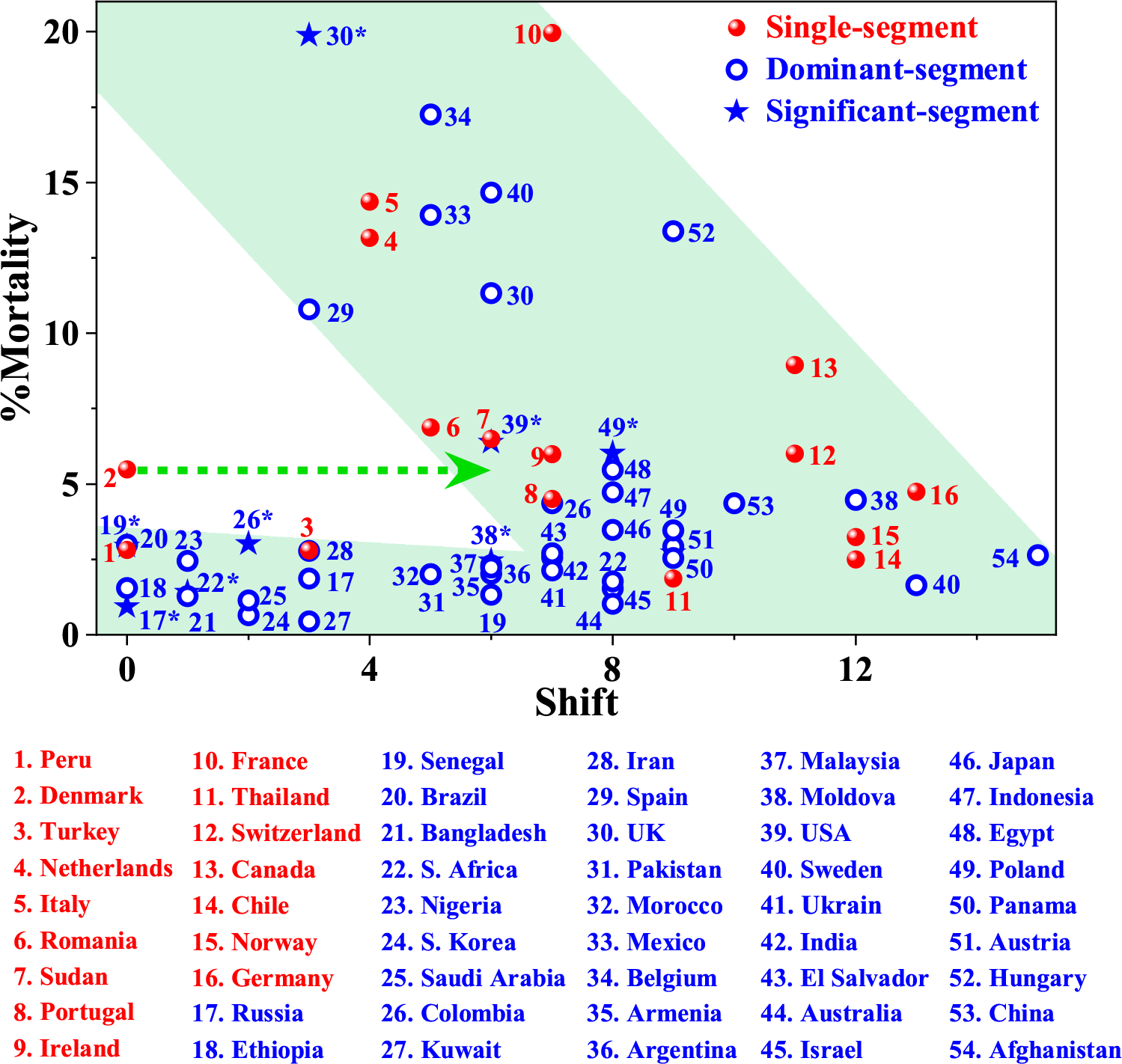
Plot of %Mortality vs. Shift obtained from scaling analysis of all countries. The red dots and blue open circles represent the countries with single-segment analysis and the dominant segment of countries with multi-segment analysis, respectively. Minor segments with infections within 80% of that in the dominant segment are also shown with stars, wherever such a situation has been encountered in our analysis.

We note that for Denmark to belong together with all other European countries with its estimated mortality rate of ⩾ 5%, its *S* parameter should be ≥5 instead of being 0, as indicated by the dashed arrow in Fig. 5. While Denmark remains a distinct exception, discussed also later in the text, the remaining countries, including all other developed countries appearing in this range marked by the inclined region, lend credence to the hypothesis that a larger shift provides a distinct advantage in keeping down the mortality rate in any given scenario. Of course, the substantial variation in the mortality rate for any given shift, as obvious in Fig. 5, also indicates that the mortality rate is not only controlled by the single parameter, *S*, and that there must be several other factors that control the mortality rate. There are huge differences between different countries in terms of the investment in the per capita health care system, there are also differences in defining what constitutes a COVID-19 related death between different countries, the accuracy of reporting is also not expected to be uniform across all nations. Therefore, it is not surprising to find that there is indeed dispersion in the mortality rate for a given value of the *S* parameter, but our analysis spanning 54 countries suggests that the Shift does have a strong influence on *M*, indicating that national efforts should be aimed at early detections of the infection, thereby giving a chance to control the mortality. A large value of the Shift will also help the nation to respond better to the anticipated demand on the health care facilities and equipment, as it gives more time to consolidate resources to any anticipated surges in demand for critical care equipment.

The early detection of infection can only be achieved by increasing the number of tests being carried out in any country along with other social aspects of awareness and reporting habits of the general population. However, increasing the number of tests has many implications in terms of the financial burden, the effort of organising large-scale testing, and availability of testing infrastructures. Given the rapid surge of the pandemic, every country must optimise its testing protocol and the number of tests to be carried out. How does a country decide what number of tests is a reasonable number? In general, every country carries out testing of anyone with a certain level of symptoms, countries differing from each other in the definition of the eligibility in terms of the severity of these symptoms. Certain countries have active policies of contact tracing, covering primary and secondary contacts; in some of these countries, the testing may be extended to cover such contacts as well. The frequency or the definition of the test eligibility may also change with changing policies and strategies within a country based on the stage of the pandemic. At times, the number of tests is expressed in terms of number of tests per million of residents of the country. Since this number necessarily only grows with time, it cannot directly reflect the course of the pandemic. On the other hand, average number of infections confirmed per unit of tests, often expressed as a percentage, and termed the %Positive cases, is a useful indicator for marking the progress of the pandemic and also for any decision-making process. For example, an increase in the number of confirmed infections per 100 tests would indicate an acceleration of the pandemic and suggest the need to increase the number of tests further. Based on these considerations, we plot the Shifts, obtained from our scaling analysis, for all segments of all countries vs. the corresponding %Positive cases for that segment, wherever the test data are available in Fig. 6(a). This plot makes it clear that there is an inverse relationship between the Shift and the %Positive cases. This correlation suggests that an effective way to build in a large shift between the detection of the infection and the mortality and consequently, reduce mortality itself (see Fig. 5), this being the primary concern of any society, is to monitor the number of infected cases per unit of tests carried out. Any acceleration of the pandemic will tend to an increase in the %Positive cases; our analysis suggests that, under such circumstances, the testing efforts in that group of population will have to be correspondingly increased in order to retain the same Shift value. This also allows one to rationalise the distribution of tests to be carried out across the nation in terms of local %Positive cases within the country as well as aiming at similar Shift and Factor that scale the mortality data on to the infection data, helping the available resources to be strategically utilised under optimal conditions.

**Figure 6:**
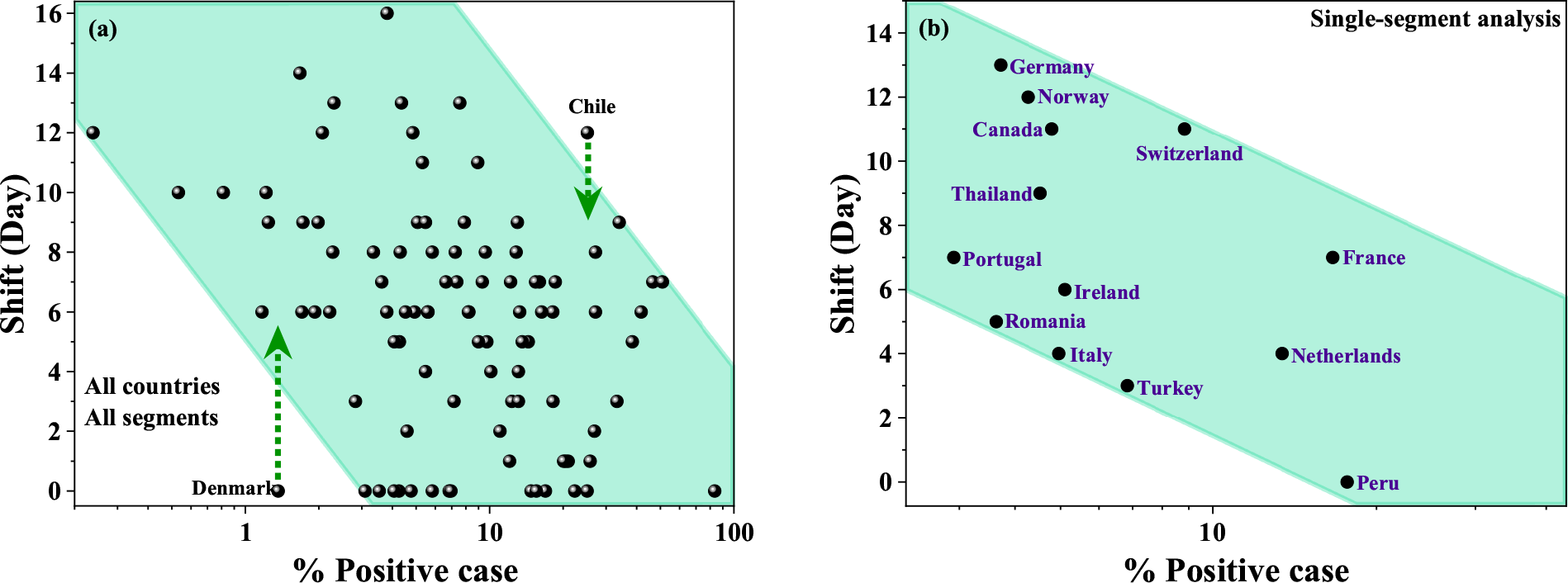
(a) Plot of Shift, obtained from the scaling behaviours of all countries and over all segments, vs. number of positive cases of infections detected for every 100 tests in that country during the corresponding period. (b) Same as in panel (a) but only for countries that could be analysed within a single segment for the entire period of analysis. This plot excludes the anomalous cases of Denmark and Chile, shown in panel (a).

Chile appears to have a slightly larger shift (12) than should be expected based on the correlation in Fig. 6(a). We note that a modest change of the Shift by 2 bringing it to 10, would have brought Chile within the shaded region as shown in the figure, suggesting that it may well be within the uncertainties discussed before. However, Denmark clearly stands out as an exception to the correlation between Shift and %positive cases. Despite this anomaly, it is interesting to note that the lowest Shift that will be compatible with Denmark’s low %Positive cases is 5 as shown in Fig. 6(a) with a dashed arrow. This expected shift of at least 5 was also shown to be anticipated based on the ∼ 5% mortality rate of Denmark in connection with our discussion of Fig. 5 correlating Shift and Mortality. In this sense, Denmark shows an underlying compatibility between its relatively low mortality rate within European countries connected with its very small %Positive cases due to a large testing program despite an anomalously small Shift value, prompting one to wonder if the death reporting in Denmark follows a different protocol compared to other countries considered here.

In summary, we have analysed the data pertaining to COVID-19 infections and deaths for 54 countries over wide periods covering the most rapid spreads of the infection and recoveries, wherever achieved. With proper segmenting of the data into different date ranges, we show that the cumulative deaths and infections as a function of time can be mapped on to each other for any given country with a shift of one with respect to the other and a normalising factor; this is achieved with minimum segments for each country, with 16 countries requiring a single segment covering the entire time range, while the remaining ones required 2 (for 14 countries) and 3 (for 24 countries) segments. The normalising factors provide reliable estimates of the mortality within the corresponding range of dates. These shifts and mortality rates are found to be spread over 0-16 days and 0.45-19.96%, respectively, indicating wide variabilities between countries as well as across different segments of individual countries requiring multi-segment analysis. Correlation of these shifts and mortalities over all segments and countries show an inverse relationship, suggesting early detection is crucial in controlling the mortality rate. Analysing further the pattern of testing of the infection in different countries during different periods, we find that %Positive cases detected through such testing correlates inversely with shifts. These results help in anticipating the progression of the pandemic and therefore, help in managing the medical and other resources in controlling along with ways to influence the mortality rate.

## Data Availability

All the data related to this study are available from the corresponding author upon reasonable request.

## Abbreviations

Following are the abbreviations used in Table 1:

*Period*: The total period investigated for any given country
*M*_*bound*_: Lowest bound on the %mortality per detected case as on the last date of analysis, defined as the total number of death divided by the total number of cases on the last date of the analysis period
*N*_*seg*_: Number of segments required for the analysis of this country
*Start date*: The first date included in the analysis of that segment
*End date*: The last date included in the analysis of that segment
*S*: Shift required for the best fit
*F*: Factor required for the best fit
*M*: Mortality rate in %, defined as (100*/F*)
*N*_*infections*_: Total number of infections in the specified period between the start and end dates
*N*_*tests*_: Total number of tests in the specified period between the start and end dates
%*Positive cases*: Number of infections detected per hundred tests carried out in the specified period between the start and end dates

## Acknowledgement

The authors thank Science and Engineering Research Board and the Department of Science and Technology, Government of India as well as Jamsetji Tata Trust for their continued support.

## Footnotes

1 It is to be noted that in all our analyses, each segment is treated independently with the assumption that the first date of the segment is the beginning of the infection; this is easily achieved by subtracting from the cumulative number of infections and deaths within the specific period under analysis the number of infections and deaths already reported prior to this period, but adding this number back for the purpose of plotting all segments together.

## Notes

### Competing Interest Statement

The authors have declared no competing interest.

### Author Declarations

The article focuses on the statistical analysis of the progression of the COVID-19 pandemic and the correlation between the infection and mortality with the help of simple scaling behavior. In view of that, the given manuscript does not seek any IRB approval.

